# Association Between Major Adverse Cardiac Events and Hormone Therapy Usage in Prostate Cancer Patients of a Diverse Cohort

**DOI:** 10.1101/2024.10.09.24314666

**Authors:** Yuanchu J Yang, Chenjie Zeng, Kerry R Schaffer, Tam C Tran, Peter J Sauer, Lincoln A Brown, Ben H Park, Joshua C Denny

## Abstract

**Importance:** Hormone therapy (HT) has led to improved overall survival for prostate cancer patients, but may also increase cardiovascular risk.

**Objective:** To compare time-to-event for major adverse cardiovascular events (MACE) between those with and without HT use in prostate cancer patients.

**Design, Setting, and Participants:** This retrospective cohort study examined 5,156 participants from the *All of Us* Research Program who were diagnosed with prostate cancer and either treated or not treated with HT (defined as exposure to a GnRH agonist, GnRH antagonist, and/or anti-androgens). Time to MACE was defined using longitudinal electronic health record data. We evaluated whether HT use affected the risk of MACE using Cox regression adjusted for established cardiovascular risk factors.

**Exposures:** HT treatment (HT-treated study group), non-HT treatment (control group without HT but with surgery, radiation treatment, and/or non-HT medical therapy), or no treatment (active surveillance control group).

**Main Outcomes and Measures:** Time-to-event for MACE, which is defined as the interval between the start of treatment (or first prostate cancer diagnosis for the no treatment group) and the date of MACE. Participants who did not develop a MACE were right censored at their last healthcare provider visit.

**Results:** The final cohort included 5,156 participants; 851 in the HT treatment group, 624 in the non-HT treatment group, and 3,681 in the no treatment group. In participants with pre-treatment dyslipidemia, HT was found to be associated increased risk of MACE (HR, 1.52; 95% CI, 1.19-1.95; P <.001), while in those without pre-existing dyslipidemia, no association were found (HR, 0.96; 95% CI, 0.71-1.30; P = .81). Similar patterns were found across race and ethnicity groups. The combined androgen blockade was statistically significantly associated with MACE in participants with pre-existing dyslipidemia (HR, 1.58; 95% CI, 1.13-2.19; P= .006) and no association in participants without pre-existing dyslipidemia (HR, 0.96; 95% CI, 0.71-1.30; P= .81). We also observed that HT was associated with prolongation of the QTc interval (P= .02).

**Conclusions and Relevance:** HT treatment was associated with an increased risk for MACE participants with pre-existing dyslipidemia. These results suggest that increased risk stratification can help improve CV outcomes when deciding treatment regimens.

## Introduction

Prostate cancer (PCa) is the most common non-cutaneous malignancy in men in the United States, ^1^ with over 200,000 new cases and 30,000 deaths annually. ^2^ Androgens drive PCa progression, ^3^ ^4^ making hormone therapy (HT) a cornerstone of treatment for localized and advanced disease. ^5^ ^6^ However, HT is associated with metabolic risks, including lipid dysregulation, insulin resistance, and increased blood pressure. ^7^ ^8^ ^9^ ^10^ ^11^ ^12^ Given the role of androgens on cardiac myocyte function, HT has also been associated with QTc interval prolongation and QRS interval shortening. ^13^ ^14^

Since 2006, the link between HT and major adverse cardiac events (MACE) has been consistently supported by observational studies. ^15^ ^16^ ^17^ ^18^ However, randomized controlled trials (RCTs) show conflicting results. While the phase III RTOG 85-31 trial found no increased CV mortality with HT, ^19^ a meta-analysis of five RCTs showed a significant rise in cardiac events. ^20^ Moreover, evidence suggests that cardiovascular disease (CVD) risk varies by HT type; for example, GnRH antagonists seem to pose a lower risk than GnRH agonists ^21^ and adding anti-androgens (abiraterone, first-generation androgen antagonist, and second-generation androgen antagonist) to a HT regimen appears to further increase CV risk. ^22^

CVD is the most common non-cancer cause of death among PCa patients. ^23^ ^24^ As both life expectancy and HT duration continue to increase, CV risk has become more prominent in PCa patients. This risk has prompted caution from the American Heart Association, American Cancer Society, and American Urological Association on the use of HT. ^25^ Despite this, no standardized guidelines exist for assessing MACE risk associated with HT. Additionally, the role of common CV risk factors like dyslipidemia, hypertension, and type 2 diabetes in HT-induced MACE remains unclear. ^26^ ^27^ ^28^ ^29^ ^30^ ^31^ ^32^ The lack of racial and geographic diversity in RCTs ^33^ ^34^ ^35^ further complicates applying these findings to the general population, highlighting the need for better risk stratification and more inclusive research.

*All of Us* is an actively enrolling, national cohort study. Currently, over 400,000 participants are enrolled, with more than 80% of this population comprised of individuals underrepresented in biomedical research. ^36^ To better account for real-world patient diversity and treatment patterns, we sought to further understand the risk of MACE with HT usage by studying participants in the *All of Us* Research Program.

## Methods

### Study Design

This retrospective cohort study utilized the Controlled Tier Dataset v7 available from participants enrolled in the *All of Us* Research Program. Data analysis was conducted from February 10, 2024, to August 17, 2024. Participants enrolled in the *All of Us* Research Program granted consent for their deidentified electronic health record to be used for research, and usage was determined by the *All of Us* Research program institutional review board to be non-human subject research. This study followed the Strengthening the Reporting of Observational Studies in Epidemiology (STROBE) reporting guidelines. ^37^

### Study Population

We identified 6,120 participants with an International Classification of Diseases (ICD) Ninth Revision (ICD-9) code of 185 or an ICD-10 code of C61, which represents a diagnosis of “Malignant Neoplasm of the Prostate.” These 6,120 participants were categorized into three groups: HT treatment, non-HT treatment, and no treatment. Participants in the HT treatment group had their first exposure to a HT after the appearance of a PCa ICD code. Participants in the non-HT treatment group were not exposed to HT, but received either surgical therapy, radiation therapy, or non-HT medical therapy after their first documented PCa diagnosis (Supplemental Table 1). Participants in the no treatment group did not have any previously defined PCa therapies after their first PCa diagnosis date, which was suggestive of patients who were under active surveillance for their PCa. Index date for participants in the HT treatment group and non-HT treatment group were defined as the earliest date of HT exposure and non-HT PCa treatment, respectively. Participants in the no treatment group had their index date set to the first date of their PCa diagnosis. We excluded from analysis 845 participants with PCa who had their first time MACE before or on their index date.

### Exposures

The primary exposure of our study was treatment with HT. We additionally stratified HT treatment by regimens. A HT drug was considered to be included in a regimen if a participant had exposure to the drug after PCa diagnosis (Full list of HT drugs available in Supplemental 1). HT regimens included in our analysis were: GnRH agonist only, GnRH antagonist only, GnRH agonist and antagonist, combined androgen blockage (CAB), and anti-androgen only. Participants in the GnRH agonist only and GnRH antagonist only regimens received only their aforementioned HT. Participants in the GnRH agonist and antagonist group have a history of receiving both GnRH agonists and GnRH antagonists, but no anti-androgen drugs. Participants in the CAB regimen received a GnRH agonist and/or a GnRH antagonist, in addition to an anti-androgen. Participants in the anti-androgen only group received exclusively anti-androgens as part of their treatment regimen.

Our non-HT treatment control group participants were exposed to non-HT medical therapy, surgery, and/or radiation for their PCa. Within this group, 30 participants had non-HT medical therapy, 262 had surgery, and 336 had radiation as their first treatment.

### Outcomes

The primary outcome measured was MACE. The time-to-event was defined as the interval from the index date until the date of first MACE. We defined MACE as the appearance of an ICD-9/ICD-10 diagnosis code for myocardial infarction, stroke, or heart failure (Supplemental Table 2). Participants who did not develop a MACE were right censored at their last visit to a healthcare provider. We compared the time-to-event between the HT, non-HT, and no treatment groups.

Secondary outcomes were the change in QRS, QTc, and PR intervals after index date. Within our final cohort, we utilized measured electrocardiogram (ECG) intervals before and after a participant’s index date. To be most representative of a patient’s baseline physiology, we removed interval values that indicated acute pathologies (QTc > 500 milliseconds, QRS > 120 milliseconds, PR > 220 milliseconds). Following this, we calculated the difference between the most recent interval after a participant’s index date and that of the most recent ECG reading before. We compared the intervals between the HT treatment group with the combined non-HT treatment and no treatment groups.

### Covariate Measurement

Covariates included in the primary Cox regression model were selected on the basis of known cardiac risk. Covariates used in our primary analysis were age at index date, dyslipidemia, type 2 diabetes, hypertension, chronic kidney disease, peripheral vascular disease, statin usage, self-reported race/ethnicity, and smoking history (Supplemental Table 3). Dyslipidemia, type 2 diabetes, hypertension, chronic kidney disease, and peripheral vascular disease were defined as the presence of an ICD-9/ICD-10 diagnosis code on or before a participant’s index date. Statin usage was defined as the presence of a statin drug exposure on or before a participant’s index date (Supplemental Table 1). Participant self-reported race, ethnicity, and smoking history was ascertained through the *All of Us* survey data.

We additionally included pre-treatment metastasis status as a covariate in a subgroup analysis of the HT and non-HT treatment groups. Metastasis status was predicted as the first-time appearance of ICD-9/ICD-10 diagnosis code of “secondary malignancy” (Supplemental Table 3) after the first ICD-9/ICD-10 PCa diagnosis code and before a participant’s index date. We additionally included pre-treatment metastasis status as a covariate in a subgroup analysis of the HT and non-HT treatment groups. Metastasis status was defined as the first-time appearance of ICD-9/ICD-10 diagnosis code of “secondary malignancy” (Supplemental Table 3) after first ICD-9/ICD-10 PCa diagnosis code and before a participant’s index date. This approach is similar to previous methods used to identify metastatic PCa using structured data in EHR. ^38^

### Statistical Analyses

Time-to-event cumulative incidence curves were utilized to visualize a participant’s MACE date from their index date, and results between treatment groups were compared using log-rank tests. To evaluate the association of potential risk factors, we utilized a Cox proportional hazards model adjusting for covariates. Pre-planned interactions terms analyzed corresponded to the effects of HT on baseline physiology, and were HT × Type 2 Diabetes, HT × Hypertension, and HT × Dyslipidemia.

We utilized unpaired student’s t-test to compare between the change in ECG intervals from different treatment groups. P-value for interactions was derived from the likelihood ratio test, comparing the Cox model without the interaction to the Cox model with the interaction. All reported P-values are two-sided, and the statistical significance threshold was predetermined at .05. Cumulative incidence curve visualization and corresponding log-rank test was conducted using ggsurvplot and ggplot2 packages. Cox proportional hazards models were built using the R survival package. All data analyses were conducted using R version 4.4.0 in the *All of Us* Researcher Workbench.

## Results

### Baseline Characteristics

Our final study cohort included 5,156 participants: 851 participants in the HT treatment group, 624 participants in the non-HT treatment group, and 3,681 participants in the no treatment group. Baseline characteristics and demographic information for the cohort are shown in Table 1. In the HT treatment group, 136 (16.0%) participants developed a MACE after their index date. In the non-HT treatment group and the no treatment group, 128 (20.5%) and 581 (15.8%) participants respectively developed a MACE. Mean electronic health record follow up after index date was 4.2 years for the HT treatment group, 7.3 years for the non-HT treatment group, and 6.1 years for the no treatment group. A total of 719 (13.9%) of our cohort self-identified as Black/African-American and 383 (7.4%) self-identified as Hispanic/Latino.

**Table 1.**
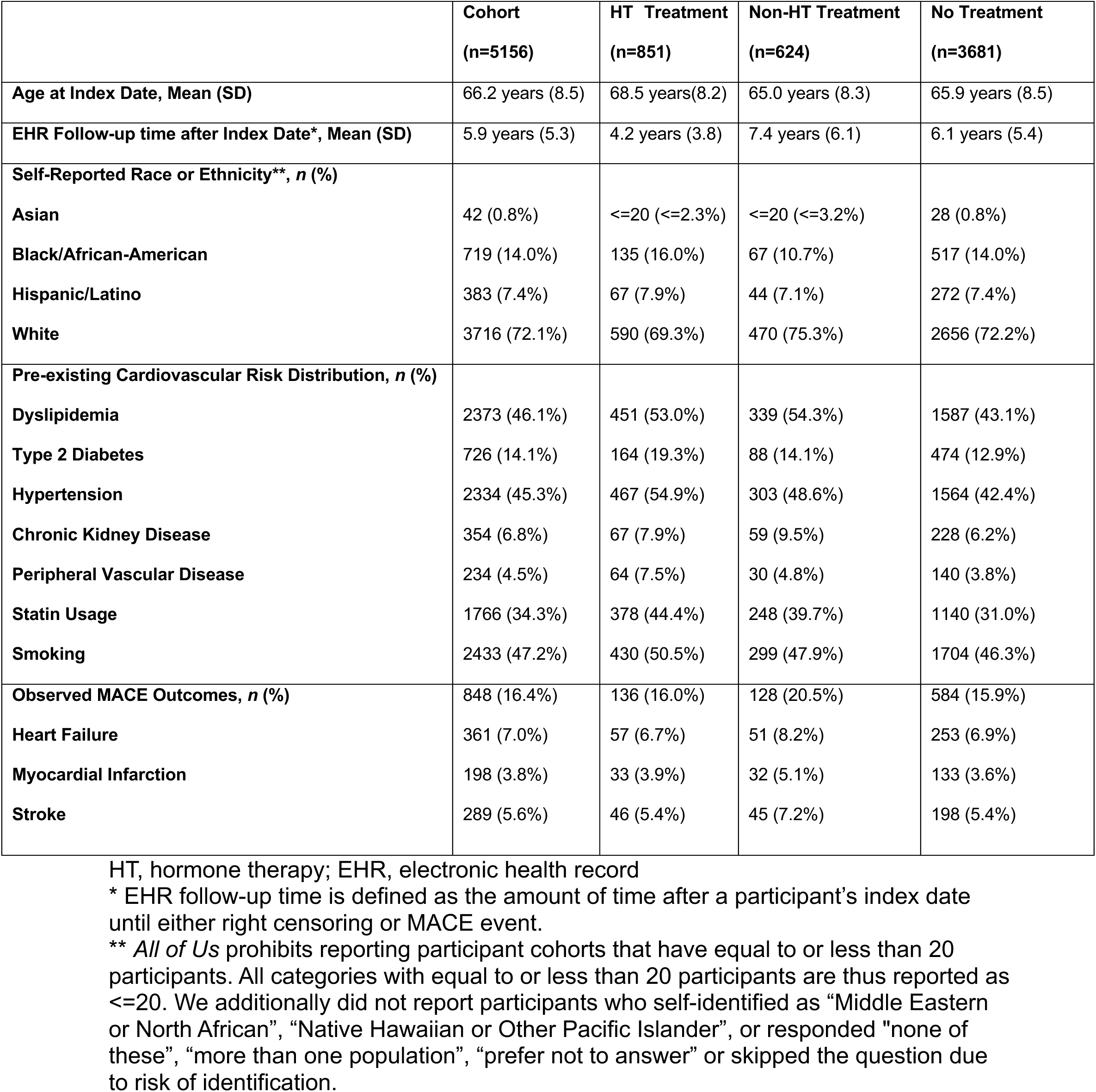
Baseline Characteristics of Study Cohort.

### MACE Risk after HT Treatment

Unadjusted cumulative incidence curves (Figure 2) showed that participants in the HT treatment group had lower time-to-event than either the non-HT treatment group (log rank test; P= .004) or the non-treatment group (log rank test; P < .001). There was no difference between the non-HT treatment group and the no treatment group (log rank test; P=.57). Because of this, we combined the non-HT treatment group and no treatment groups into our reference group for our Cox proportional hazard models (Supplemental Table 4). In an unadjusted, univariate Cox regression, HT treatment was associated with decreased MACE time-to-event (HR, 1.45; 95% CI, 1.20-1.75; P < .001). After adjustment for baseline cardiovascular covariates in our multivariate analysis, HT treatment remained associated with lower MACE time-to-event (HR, 1.22; 95% CI, 1.01-1.48; P = .03). Independent risk factors for MACE time-to-event were pre-existing type 2 diabetes, hypertension, chronic kidney disease, age, and smoking history. Statin usage was associated with a protective effect. Self-identification as Black/African-American (HR, 1.29; 95% CI, 1.04-1.58; P = .02) and as Hispanic/Latino (HR, 1.51; 95% CI, 1.16-1.96; P = .002) was found to be associated with MACE time-to-event (Supplemental Table 4). Multivariable Cox model of the Black/African-American or Hispanic/Latino participant subgroups are shown in Supplemental Table 7.

**Figure 1.**
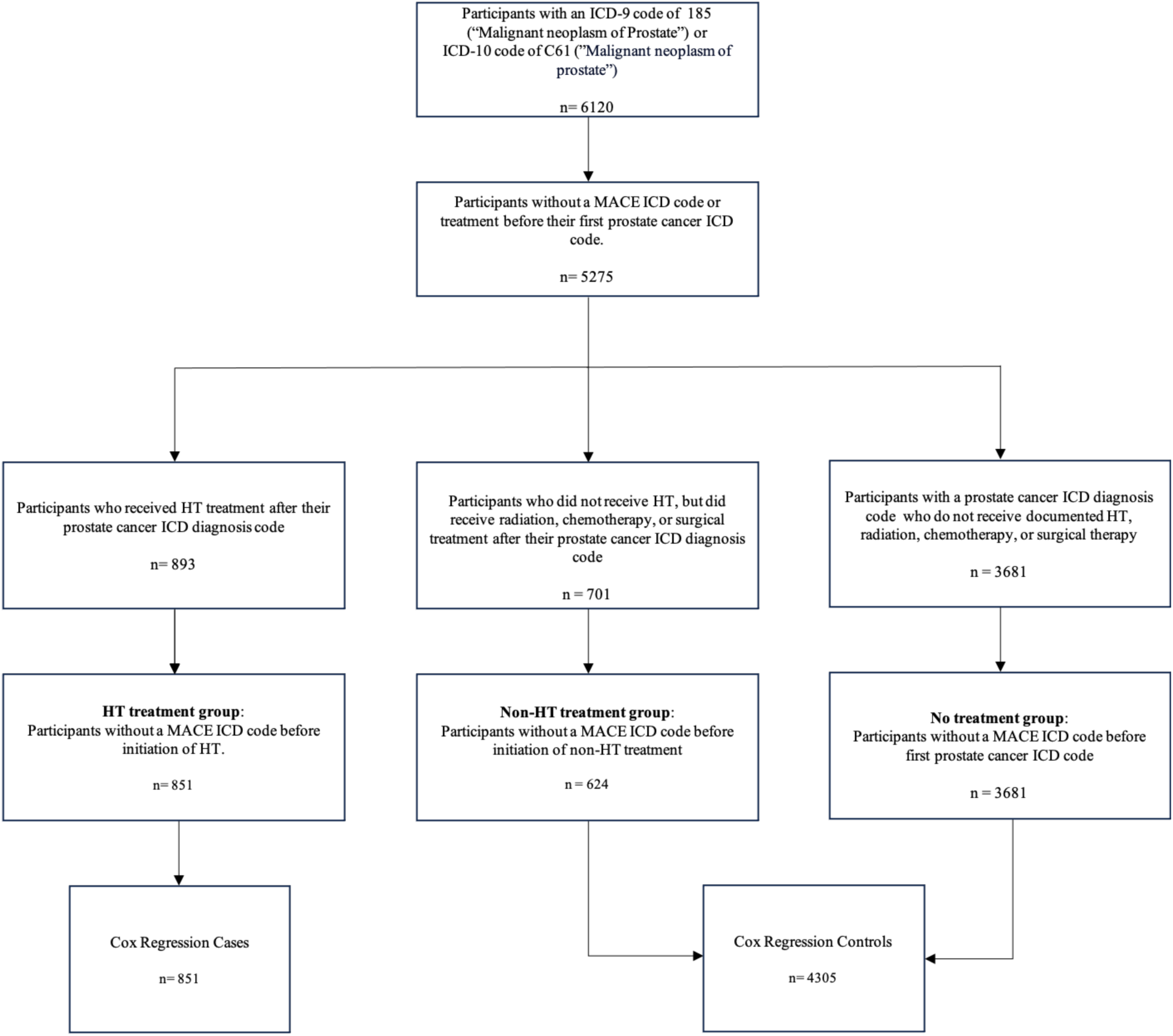
Study Flowchart.

**Figure 2.**
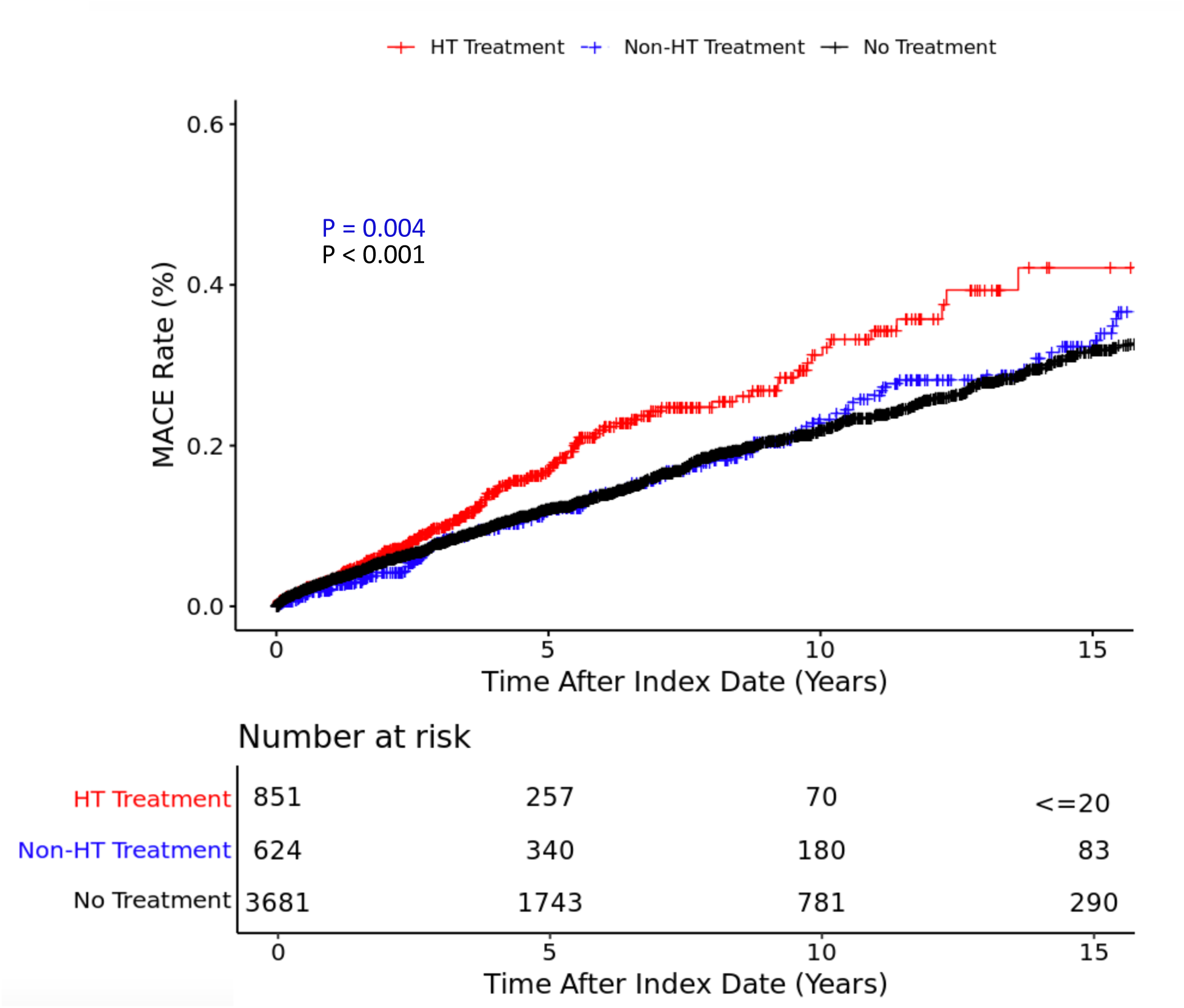
Cumulative Incidence Plot by Treatment Groups. P values indicate the log rank test between the HT Treatment Group and either the non-HT treatment (blue) or no treatment (black) groups. *\*All of Us* prohibits reporting participant cohorts that have equal to or less than 20 participants. All categories with equal to or less than 20 participants are thus reported as <=20.

### HT Interaction with Dyslipidemia

We found the HT and dyslipidemia interaction term was significant in our multivariate analysis (HR, 1.57; 95% CI, 1.03-2.34; P = .03), which suggested a synergistic effect on MACE risk between HT and pre-treatment dyslipidemia. No other pre-determined interaction terms were found to be significant (Supplemental Table 5). As a result, we stratified our overall Cox model based on participant baseline dyslipidemia status (Table 2). In participants with pre-existing dyslipidemia, HT treatment was associated with decreased MACE time-to-event (HR, 1.52; 95% CI, 1.19-1.95; P <.001). In participants without pre-existing dyslipidemia, HT treatment had no association with MACE time-to-event (HR, 0.96; 95% CI, 0.71-1.30; P = .81).

**Table 2.** Cox Model for MACE Time-To-Event by Pre-treatment Dyslipidemia.

|  | Participants with Pre-existing dyslipidemia |  | Participants without Pre-existing dyslipidemia |  |
| --- | --- | --- | --- | --- |
|  | Hazard Ratio (95% CI) | P-Value | Hazard Ratio (95% CI) | P-Value |
| **HT** | 1.52 (1.19-1.95) | <.001 | 0.96 (0.71-1.30) | .81 |

HT, hormone therapy

### Stratification by HT Regimen

Next, we performed analyses after stratifying by HT regimen. In a multivariable-adjusted Cox regression model, the CAB regimen was associated with decreased MACE time-to-event in the overall cohort (HR, 1.31; 95% CI, 1.03-1.68; P= 0.03). No other treatment regimen was found to be associated with MACE time-to-event in our overall cohort. In participants with pre-existing dyslipidemia (Table 3), the CAB regimen showed increased risk of MACE (HR, 1.52; 95% CI, 1.19-1.95; P< .001). No association was appreciated between the CAB regimen and MACE time-to-event in participants without pre-existing dyslipidemia (HR, 0.96; 95% CI, .71-1.30; P= .81). Similar associations were seen when anti-androgen drugs in the CAB regimen were limited to abiraterone or second-generation androgen antagonists (Supplemental Table 6).

**Table 3.** Cox Model for MACE Time-To-Event stratified by HT Regimen.

|  | Participants with Pre-existing dyslipidemia<br>(n=2377) |  | Participants without Pre-existing dyslipidemia<br>(n=2779) |  |
| --- | --- | --- | --- | --- |
|  | HR (95% CI) | P-Value | HR (95% CI) | P-Value |
| **GnRH Agonist Only (n<=20)** | 1.51 (0.21-10.80) | .68 | 1.46 (N/A) | .99 |
| **GnRH Antagonist Only (n=260)** | 1.51 (1.02-2.22) | .038 | .71 (0.38-1.34) | .27 |
| **GnRH Agonist and Antagonist (n<=20)\*** | 4.39 (0.61-31.6) | .14 | 1.45 (0.20-10.4) | .72 |
| **Combined Androgen Blockade (n=n409)\*\*** | 1.58 (1.14-2.19) | .006 | 1.08 (0.74-1.58) | .70 |
| **Anti-Androgen only (n=152)** | 1.34 (0.77-2.35) | .31 | 1.09 (0.57-2.05) | .84 |
| **No HT (n=4305)** | Reference | Reference | Reference | Reference |

N/A , not available due to small sample size; HT, Hormone Therapy. HR: hazard ratio.
\* Participants in the GnRH agonist and antagonist group had both exposures to GnRH agonists and GnRH antagonists after their first prostate cancer ICD code. This is likely due the result of the participant switching between hormone therapy regimens.
\*\* Participants in the combined androgen blockage group received either a GnRH agonist or GnRH antagonist, in addition to an anti-androgen after their first prostate cancer ICD code.

### Pre-treatment Metastasis in HT and non-HT Treatment Groups

Additionally, we assessed the effects of pre-treatment metastatic disease in a Cox multivariable regression model of the combined HT and non-HT treatment group (Supplemental 8). In the HT treatment group, 158 participants had metastatic disease before their index date. In the non-HT treatment group, 32 had metastatic disease before their index date. The presence of metastatic disease was associated with decreased MACE time-to-event (HR, 1.82; 95% CI, 1.23-2.71; P= .003). The aforementioned HT and dyslipidemia interaction term continued to be significant in this sub-analysis (HR, 1.81; 95% CI, 1.06-3.08; P= .03).

### Electrocardiogram Intervals

Within our cohort, there were 127 participants with measured QTc intervals, 122 participants with measured QRS intervals, and 108 participants with measured PR intervals both before and after their index date (Supplemental Table 9). We observed a significant increase in measured QTc intervals after index date when comparing the HT treatment participants to the combined non-HT treatment and no treatment participants (P= .02). No differences were observed in the QRS and PR intervals. No participant in our cohort had an ICD-9/ICD-10 diagnosis code for torsades de pointes.

## Discussion

MACE are the most common causes of non-cancer mortality within the PCa. ^23^ ^24^ Leveraging data from the *All of Us* cohort, we showed that treatment with HT is associated with a higher risk of MACE in PCa patients with pre-existing dyslipidemia. Our data also suggests that the usage of CAB may have higher CV risk than other HT regimens, especially in patients with dyslipidemia. We found that these patterns were consistent across race and ethnicity groups. In addition, we also found an association of HT with QTc prolongation. ^13^ ^14^

Currently, there exists no standardized guidelines in assessing MACE risk for patients who decide to start HT treatment. Furthermore, because of the complexity of CV disease drivers and the large number of PCa therapies available, there are questions about how to implement changes from observed associations into the real-world clinical setting. Previous studies have demonstrated that HT use is associated with an increased risk of mortality in patients with high CV disease risk, but not among those with low risk. ^21^ Our findings are consistent with these observations. Our results point specifically towards a diagnosis of dyslipidemia as a modifiable risk factor for the CV risk associated with HT, which we hypothesize is due to a combinatory effect on atherosclerosis. ^39^ ^40^ ^41^ This finding suggests areas for future research; for example, could monitoring lipid levels when considering HT be warranted, and could intensification of lipid management alleviate some of this risk.

In our cohort, though noting the smaller sample size, we observed that CAB usage was associated with greater MACE risk. The cardiotoxicity of CAB has been previously noted in randomized controlled trials. ^42^ One posited explanation is that CAB regimens have greater potency when compared to other regimens, and thus lead to increased downstream toxicities associated with androgen deprivation. After limiting the anti-androgen drugs in the CAB regimen to only second-generation androgen antagonists and abiraterone, our results remain congruent with recent studies detailing the CV risk posed by these specific anti-androgen drugs (Supplemental Table 6). ^22^

Black/African-American men are known to have a higher incidence of PCa compared to White men. ^43^ Furthermore, Black/African-American individuals suffer from greater cancer-related and CV mortality in the setting of a PCa diagnosis. ^43^ ^44^ ^45^ On the other hand, Hispanic/Latino individuals with PCa are suggested to be at decreased risk of CV mortality in this context. ^46^ After adjusting for baseline CV covariates, our analysis indicates that both Black/African-American and Hispanic/Latino participants in our cohort were at increased risk of MACE. As a whole, these results show the importance of continuing work to increase participant diversity in observational and prospective trials of PCa patients and highlights a need to further clarify if race and ethnicity should be taken into consideration clinically when assessing for CV risk.

The electrophysiological effect of HT can manifest itself as a prolonged QTc interval. ^13^ ^14^ Our study replicates previous findings of QTc prolongation. Given our results as well as that of others, routine ECG monitoring may be warranted in at-risk patients, especially if other therapies that prolong QTc are being introduced to the patient.

### Limitations

Limitations of this study include its retrospective nature, low power to meaningfully evaluate mortality, and the lack of initial cancer staging or Gleason score data available. We were also unable to perform some race/ethnicity group specific analysis due to low statistical power.

Currently, death data in the *All of Us* program were extracted from EHRs. If death were not recorded in the EHR, this data would be missing. Due to this, we are not able to perform analyses that included participant mortality.

Cancer staging and Gleason score are significantly correlated with all-cause and cancer-specific mortality in PCa patients. ^48^ ^49^ ^50^ ^51^ It is unknown if these factors directly affect cardiac health. Metastatic PCa is associated with greater CV mortality, ^23^ ^52^ but the causal relationship is unknown due to the presence of multiple confounding variables that include HT usage. After adjusting for metastasis status in our HT and non-HT treatment groups, our study is concurrent with this observation. In this sub-analysis, we consistently see a significant interaction between dyslipidemia and HT usage. Future releases of more unstructured EHR data in *All of Us* can be used to identify cancer stage. Because of this, we believe that future studies are warranted to explore the complex interplay between of Gleason scores/cancer staging and CV outcomes in PCa patients.

## Conclusions

Our analysis of the *All of Us* Research Program provides evidence that HT increases the risk of MACE in PCa patients, with this risk interacting with pre-treatment dyslipidemia. These findings provide insight that when initiating HT in patients, baseline risk stratification may help improve cardiovascular outcomes.

## Data Availability

Data from participants in the All of Us cohort is available through approved access from the All of Us researcher workbench.

https://workbench.researchallofus.org/

## Acknowledgement

We thank all participants enrolled in the *All of Us* Research Program for their trust and partnership, without which this research would not have been possible.

