## Supplementals for "Association Between Major Adverse Cardiac Events and Hormone Therapy Usage in Prostate Cancer Patients of a Diverse Cohort"

**Supplemental Table 1. List of Names within Treatment Class**

| **Categories** | **Treatment Names** |
| --- | --- |
| **GnRH Agonist** | Leuprolide,Goserelin,Histrelin,Triplorelin |
| **GnRH Antagonist** | Degarelix,Relugolix |
| **Abiraterone** | Abiraterone |
| **First Generation Androgen Antagonists** | Flutamide, Bicalutamide, Nilutamide |
| **Second Generation Androgen Antagonists** | Darolutamide, Apalutamide, Enzalutamide |
| **Non-HT Medical Therapy** | Docetaxel, Cabazitaxel, Carboplatin, Mitoxantrone, Estramustine, Sipuleucel-T, Pembrolizumab, Lynparza, Olaparib,Talazoparib, Niraparib |
| **Surgery** | Prostatectomy, Prostate Resection |
| **Radiation** | Brachytherapy, Intensity-modulated radiation therapy, External-beam radiation therapy, Image-guided radiotherapy, Stereotactic radiotherapy, Radiation therapy (NOS) |
| **Statin** | Atorvastatin, Cerivastatin, Lovastatin,Mevastatin, Pitavastatin, Pravastatin, Rosuvastatin,Simvastatin |

**Supplemental Table 2. ICD Codes for Major Adverse Cardiac Events**

| **Major Adverse Cardiac Event** | **ICD-9 Codes** | **ICD-10 Codes** |
| --- | --- | --- |
| **Myocardial Infarction** | 410.XX, 412.XX | I21.XX, I22.XX |
| **Stroke** | 430.XX, 431.XX, 432.XX, 433.XX, 434.XX | I60.XX, I61.XX, I62.XX, I63.XX, I64.XX |
| **Heart Failure** | 428.XX | I50.XX |

X indicates wild character

| **Covariate** | **ICD-9 Codes** | **ICD-10 Codes** |
| --- | --- | --- |
| **Dyslipidemia** | 272.XX | E78.XX |
| **Type 2 Diabetes** | 250.X2 and 250.X0 | E11.XX |
| **Hypertension** | 401.XX, 402.XX, 403.XX, 404.XX, 405.XX, | I10.XX, I11.XX, I12.XX, I13.XX, I14.XX, I15.XX, I1A.XX |
| **Chronic Kidney Disease** | 585.XX | N18.XX |
| **Peripheral Vascular Disease** | 443.89, 443.9X, 440.XX | I70.XX, I73.8X, I73.9X |
| **Metastatic Disease** | 196.XX, 197.XX, 198.5X | C78.XX, C77.XX, C79.5X |

**Supplemental Table 3. ICD Codes for Covariates**

X indicates wild character

**Supplemental Table 4.  Cox Proportional Hazards Regression Model for Time-to-event for MACE**

|  | **Multivariate Cox Model** | |
| --- | --- | --- |
| **Covariate** | **Hazard Ratio (95% CI)** | **P-Value** |
| **HT** | 1.22  (1.01-1.48) | .03 |
| **Dyslipidemia** | .93  (0.78-1.10) | .40 |
| **Type 2 Diabetes** | 1.26  (1.03-1.54) | .026 |
| **Hypertension** | 1.44  (1.23-1.69) | <.001 |
| **Chronic Kidney Disease** | 1.79  (1.39-2.31) | <.001 |
| **Peripheral Vascular Disease** | 1.32  (0.96-1.79) | .08 |
| **Age (per year)** | 1.05  (1.04-1.05) | <.001 |
| **Statin Usage** | 0.79  (0.66-0.94) | .009 |
| **Smoking History** | 1.20  (1.05-1.38) | .008 |
| **Race/Ethnicity** |  |  |
| Asian (n=42) | .96  (.43-2.16) | .93 |
| Black (n=719) | 1.29  (1.04-1.58) | .02 |
| Hispanic/Latino (n = 383) | 1.51  (1.16-1.96) | .002 |
| Middle Eastern or North African (n = 22) | .66  (.21-2.07) | .47 |
| More than One Population (n=41) | 1.02  (.51-2.07) | .95 |
| Native Hawaiian or Other Pacific Islander* (n<=20) | N/A | N/A |
| Unknown** (n=232) | 1.04  (.75-1.44) | .82 |
| White (n=3716) | Reference | Reference |

N/A , not available due to small sample size ; HT, hormone therapy;

**All of Us* prohibits reporting participant cohorts that have equal to or less than 20 participants. All categories with equal to or less than 20 participants are thus reported as <=20.

**Unknown includes participants who responded "none of these”, “prefer not to answer” or skipped the question.

**Supplemental Table 5.  Cox Proportional Hazards Regression Models for Time-to-event for MACE with Interactions**

|  | **Multivariate Cox Model + Interactions Terms** | |
| --- | --- | --- |
| **Covariate** | **Hazard Ratio (95% CI)** | **P-Value** |
| **HT** | .97  (0.69-1.37) | .88 |
| **Dyslipidemia** | .84  (0.69-1.00) | .056 |
| **Type 2 Diabetes** | 1.29  (1.03-1.62) | .028 |
| **Hypertension** | 1.51  (1.27-1.81) | <.001 |
| **Chronic Kidney Disease** | 1.82  (1.41-2.35) | <.001 |
| **Peripheral Vascular Disease** | 1.36  (.99-1.83) | .051 |
| **Age (per year)** | 1.04  (1.03-1.05) | <.001 |
| **Statin Usage** | .78  (.65-0.93) | .005 |
| **Smoking History** | 1.19  (1.04-1.36) | .011 |
| **HT × Hyperlipidaemia** | 1.57  (1.03-2.34) | .03 |
| **HT × Type 2 Diabetes** | 1.03  (0.64 -1.66) | .90 |
| **HT × Hypertension** | 0.97  (0.64- 1.46) | .87 |

HT, hormone therapy

**Supplemental Table 6. Cox Proportional Hazards Regression Models stratified by HT Regimen; anti-androgens in combined androgen blockage limited to only abiraterone and second-generation androgen antagonists**

|  | **Participants with Pre-existing Dyslipidaemia** | | **Participants without Pre-existing Dyslipidaemia** | |
| --- | --- | --- | --- | --- |
|  | Hazard Ratio (95% CI) | P-Value | Hazard Ratio (95% CI) | P-Value |
| **GnRH Agonist (n<=20)** | 1.53 (.21-10.92) | .67 | 1.45 (N/A) | .99 |
| **GnRH Antagonist (n=260)** | 1.52 (1.03-2.24) | .035 | .71 (.38-1.32) | .27 |
| **GnRH Agonist and GnRH Antagonist n<=20)** | 4.48 (.62-32.0) | .14 | 1.43 (.20-10.3) | .72 |
| **Combined Androgen Blockade (Abiraterone and Second-Generation Androgen Antagonist; n=161)** | 2.84 (1.84-4.38) | <.001 | 1.17 (.67-2.04) | .59 |
| **Combined Androgen Blockade (First-Generation Androgen Antagonist;n=248)** | 1.03 (.65-1.64) | .89 | 1.02 (.62-1.69) | .95 |
| **Anti-Androgen Only (n=152)** | 1.35 (.78-2.39) | .30 | 1.07 (.57-2.02) | .84 |
| **No HT (n=4305)** | Reference | Reference | Reference | Reference |

HT, hormone therapy

**Supplemental Table 7. Cox Proportional Hazards Regression Models in Black/African-Americans and Hispanic/Latino**

| **Covariate** | **Black/African-American (n=719)** | | **Hispanic/Latino (n=383)** | |
| --- | --- | --- | --- | --- |
|  | Hazard Ratio (95% CI) | P-Value | Hazard Ratio (95% CI) | P-Value |
| **HT** | .38 (0.11-1.35) | .14 | .67 (.20-2.24) | .51 |
| **Dyslipidemia** | .56 (0.33-0.95) | .032 | .50 (.24-1.06) | .07 |
| **Type 2 Diabetes** | 1.28(.73-2.22) | .39 | 1.33 (.66-2.66) | .43 |
| **Hypertension** | 1.39 (.87-2.23) | .16 | 1.49 (.80-2.75) | .20 |
| **Chronic Kidney Disease** | 2.68 (1.59-4.51) | <.001 | 1.82 (.68-4.92) | .23 |
| **Peripheral Vascular Disease** | 1.18 (.54-2.55) | .68 | .23 (.03-1.89) | .20 |
| **Age (per year)** | 1.05 (1.02-1.07) | <.001 | 1.04 (1.01-1.07) | .01 |
| **Statin Usage** | 1.00 (.61-1.65) | .99 | .78 (.34-1.74) | .55 |
| **Smoking History** | 1.54 (1.06-2.25) | .024 | 1.48 (.90-2.45) | .14 |
| **HT ×  Dyslipidaemia** | 2.85 (.96-8.45) | .058 | .53 (.05-4.61) | .57 |
| **HT × Type 2 Diabetes** | 0.65 (.21-1.97) | .44 | .46 (.04-4.21) | .49 |
| **HT × Hypertension** | 2.74 (0.74-10.30) | .13 | 1.63 (.35-7.65) | .54 |

HT, hormone therapy

**Supplemental Table 8.  Subgroup Cox Model with Pre-treatment Metastasis Status in Combined HT and non-HT Treatment Groups (n=1475)**

|  | **Hazard Ratio (95% CI)** | **P-Value** |
| --- | --- | --- |
| **HT** | .77 (0.50-1.19) | .24 |
| **Dyslipidemia** | .77 (0.52-1.15) | .20 |
| **Type 2 Diabetes** | 1.26(0.23-2.18) | .41 |
| **Hypertension** | 1.46 (0.99-2.16) | .06 |
| **Chronic Kidney Disease** | 1.58 (1.03-2.43) | .04 |
| **Peripheral Vascular Disease** | 0.83 (0.47-1.47) | .53 |
| **Age (per year)** | 1.04 (1.02-1.06) | <.001 |
| **Statin Usage** | 0.76 (0.57-1.02) | .06 |
| **Smoking History** | 0.96 (0.75-1.23) | .74 |
| **Metastasis Status** | 1.82 (1.23-2.71) | .003 |
| **HT × Dyslipidemia** | 1.81 (1.06-3.08) | .03 |
| **HT × Type 2 Diabetes** | 1.08 (0.54-2.15) | .83 |
| **HT × Hypertension** | 1.13 (.21-1.97) | .65 |

HT, hormone therapy

**Supplemental Table 9. Electrocardiogram Interval Comparisons Between Subgroups**

| **EKG Interval** | **HT Treatment** | **Non-HT Treatment** | **No treatment** | **P- value** |
| --- | --- | --- | --- | --- |
| QTc (n=127)  Pre-index Interval  Post-index Interval  Mean Change | 427.7 ms  442.0 ms  +14.2 ms | 405 ms  408 ms  +3 ms | 424.0 ms  428.0 ms  +4.0 ms | .02 |
| QRS (n=122)  Pre-index  Post-index  Mean Change | 80.8 ms  83.2 ms  +2.4 ms | 84.5 ms  91.2 ms  +6.7 ms | 77.4 ms  78.2 ms  +0.8 ms | .76 |
| PR (n=108)  Pre-index  Post-index  Mean Change | 167.6 ms  167.0 ms  -0.6 ms | 164.4 ms  166.0 ms  +1.6 ms | 166.7 ms  166.8 ms  +0.1 ms | .82 |

HT, hormone therapy; MS, milliseconds
